# Exploring the Association between Contraceptive Use and Women’s tolerance towards Domestic Violence: Evidence from MICS 2019 Survey

**DOI:** 10.1101/2023.07.15.23292717

**Authors:** Tanvir Haider, Md Fuad Al Fidah, Md Emam Hossain, Syed Nafi Mahdee, Syeda Sumaiya Efa

## Abstract

**Background:** Access to contraception is crucial for preventing unintended pregnancies and promoting reproductive rights. In Bangladesh, the contraceptive prevalence rate (CPR) has increased over the decades. However, domestic violence (DV) remains a major concern. Women who experience DV are more likely to have unintended pregnancies and abortions. They are also more likely to live in poverty and experience depression and anxiety.

**Objective:** This study aimed to assess the relationship between contraceptive use and women’s tolerance of DV in Bangladesh.

**Methods:** Data from the Bangladesh Multiple Indicator Cluster Survey (MICS) 2019 was used for the study. The sample size was 47,692 women aged 15-49; data were analyzed using Jamovi software (version 2.3.26). Ethical considerations were strictly followed. *Results*

The study found a contraception utilization rate of 66.5% among Bangladeshi women of reproductive age. Factors associated with contraceptive use included age, residence, education level of the woman, marital structure, wealth index, and tolerance for domestic violence. Women with high tolerance towards domestic violence had higher odds (AOR: 1.1; 95% CI: 1.017-1.189; p = 0.017) of using contraception than women with no tolerance.

**Conclusion:** The study’s result suggest a complex relationship between contraceptive use, women’s tolerance of domestic violence, and other factors such as age, education, wealth, location, etc. Programs and policies are needed to promote contraceptive use in Bangladesh. Efforts should target socioeconomic, cultural, and psychological factors and barriers which are faced by women exposed to domestic violence. By tackling these challenges, programs, and policies can effectively promote contraception use and improve women’s reproductive health.

## Introduction

The well-being of individuals and the community highly depends on women’s reproductive health and access to contraception.^1^ Unintended pregnancies can be prevented, leading to a greater control over the reproductive rights, if individuals gain adequate access to contraceptives (CP). Additionally, high-risk pregnancies can be reduced, and maternal and child health can be promoted by ensuring proper access to contraception.^1^ Improved gender equality and women’s education, reduced poverty, etc., are known to be associated with the use of contraception.^2^

Family planning services are crucial to achieving sustainable development goals in low- and middle-income countries, as these countries account for most maternal and newborn deaths.^3^ Family planning or contraceptive methods reduce unexpected pregnancies, unsafe abortions, HIV/AIDS, and other STDs.^4-7^ Contraception helps to achieve demographic, socioeconomic, and environmental goals, especially in nations with high fertility rates.^7^ More and more research is being conducted on family planning, showing how different factors at the individual, family, geographical, sociocultural, and healthcare system levels can affect how people either use contraceptives sub optimally or don’t use these methods effectively.

In the Southeast Asian region the fertility rate is quite high, including in Bangladesh. In this country, 16.744 births will occur per 1000 people in 2023.^8^ However, the proportion of couples using contraceptive methods, also known as the contraceptive prevalence rate (CPR), has risen dramatically in the recent years, from only 8.0% in 1975 to 62.4% in 2014.^9,10^ This rising trend in contraceptive prevalence offers many benefits. One such benefit is the declining of the total fertility rate (TFR); from 6.3 children per woman in 1975 to 2.3 in 2014.^9^ It also reduced newborn and maternal mortality rates from 88.0 deaths per 1000 live births in 1993-94 to 38.0 in 2014.^11,12^ However, CPR and TFR advancements have stagnated recently. In 2017, the CPR was 62.0%, equal to 2014, while the TFR has been 2.3 children per woman since 2011.^10,13^ In some cases, the challenges surrounding family planning are more complicated and linked to sociocultural structures, such as discrimination based on gender or domestic violence (DV).^14^ These issues may be difficult to address through typical intervention strategies as they may be deeply ingrained in society. Women frequently experience domestic violence, although men can also be victims.^15^

The mistreatment of female family members within households is not uncommon and has been happening for a long time. The types of violence, both physical and psychological, differs greatly across societies and cultures. Bangladesh is no exception, and both urban and rural women have been significantly affected by domestic violence.^16^ Over the past decade, many atrocious instances of domestic violence against women have led experts and professionals to recognize its detrimental effects on society as a whole and develop effective measures to address it. Bangladesh has one of the highest rates of violence against women globally, with 50 to 70 percent of women in the country reporting abuse by their male partners.^17^ In Bangladesh, domestic violence is a major concern affecting individuals from all walks of life in rural and urban areas. Usually, the spouse is the one who commits violence against their partner. Distressingly, this violence has become a deeply ingrained cultural norm in Bangladeshi society, handed down from generation to generation. The idea that domestic violence is more prevalent among rural women due to limited access to human rights organizations and lack of awareness of their rights, while urban women are less likely to be victims, is often discussed.^18^ However, the probability of experiencing domestic violence is equal for women residing in rural as well as urban areas in Bangladesh.^19^

Domestic violence has been linked with an increased rate of unwanted childbirth, primarily through the restrictive practice of contraception by reproductive women. Studies suggest that tolerance towards DV may account for the limitation or nonuse of contraceptive methods among women of reproductive age. However, most of these studies have been conducted in the Western world^20, 21,22^and underdeveloped nations still lag in exploring such relationships. This study aims to assess the relationship between contraceptive use and women’s tolerance of domestic violence in Bangladesh.

## Methods

### Data overview

This study used nationally representative data from the Bangladesh Multiple Indicator Cluster Survey (MICS) 2019. Enumeration areas (EAs) were chosen as the primary sampling entities for the survey’s two-stage sampling strategy. Initially, 3220 EAs were selected from 64 districts using a probability proportional to size method. In the second stage, 20 households were randomly selected from each cluster (EA), yielding a total sample size of 64,400 households, with approximately 1,000 households from each stratum. The MICS questionnaire for individual women (15-49 years) was then administered face-to-face to selected individuals in the household. All variables in this study were measured and recorded based on the responses of the individuals during interviews.^23^

Women aged 15 to 49 years were included in the present study from the survey’s publicly accessible women dataset (wm file), then relevant variables were merged from the households dataset (hh file), and household members dataset (hl file). Datasets were merged using unique identifiers following the guideline provided by MICS.^24^ A total of 47,692 women were considered for analysis after excluding individuals who did not provide explicit consent, pregnant women at the time of interview, and missing values for women on information regarding current contraceptive use, and tolerance towards domestic violence.

### Methods of contraceptive use

In our study data were collected on methods of contraceptive use by women of reproductive age. These methods included: male condoms, female condoms, diaphragm, foam or jelly, female sterilization, male sterilization, intrauterine device, injectables, implants, oral contraceptives, lactational amenorrhea method, periodic abstinence or rhythm, withdrawal, and “others.” Women who did not use any of the aforementioned methods were categorized as “No” and coded as “0”, and women who used at least one of these methods were categorized as “Yes” and were coded as “1”.

### Tolerance towards DV

Tolerance towards DV was another variable considered in our study. It was calculated using the participant’s answer to 5 questions that asked if the participant thought “a man was justified for hitting his wife” in the following situations: 1. Going out without telling him, 2. Neglecting the children, 3. Arguing with him, 4. Refusing to have sex with him, and 5. burning the food. For each question, the options were: “Yes,” “No,” and “Don’t know.” These items were recoded using 0 for “No” and “Don’t know” and 1 for “Yes.” Then all five items were added to get a variable that showed the number of reasons for DV approved by each participant, which ranged from 0 to 5. Next, this new variable was categorized using 3 categories: “No tolerance” (if no reason was approved), “Low tolerance” (if 1-3 reasons were approved), and “High tolerance” (if 4-5 reasons were approved).

### Access to media

Media access was assessed by reading newspapers, listening to the radio, and watching TV. A new variable, “Media access,” was created using the summation of 3 variables: frequency of reading newspapers or magazines, frequency of listening to the radio, and frequency of watching TV. This variable had 3 categories: Poor access (0–3 score), Moderate access (4–6 score), and Good access (7–9 score).

### Marital structure

Marital structure was considered as such: Women whose husband had more than one wife were considered as having polygamous marriage, and women whose husband had only one wife were considered as having a monogamous marriage.

## Ethical clearance

The study analyzed UNICEF survey data without disclosing information to safeguard participants’ privacy. These secondary data studies did not require ethical approval because the data set was de-identified, and authorization was obtained from the competent authority. However, the Bangladesh Bureau of Statistics and UNICEF obtained informed consent before the survey.

## Statistical analysis

The present investigation employed IBM SPSS version 27 to integrate various datasets and jamovi version 2.3.26 to conduct data analysis.^25^ We computed frequency distribution, percentage, mean, and standard deviation for descriptive statistics. Continuous data were presented as mean and standard deviation, whereas categorical variables were expressed as counts and percentages. Concerning inferential statistics, the Chi-square test was used to detect any association between categorical variables. Furthermore, we developed a logistic regression model, incorporating all significant variables identified by the chi-square tests, to assess the strength of association. The level of significance was set at p<0.05.

## Results

The procedure used to select participants for this investigation is described in detail in Table 1 shows the demographic characteristics of Bangladeshi women of reproductive age (15-49). The study enrolled a total of 47,692 individuals. Out of which, most (41.7%) belonged to 35-49 years age group, while 21.5% and 36.8% belonged to the age group 15-24 years and 25-34 years, respectively, with a mean age of 32.6 (± 8.5) years. Most of the participants (80.2%) in this study were from rural areas, while 19.8% lived in urban areas. Among the participants, 43.0% had secondary education, 26.0%, 18.5%, and 12.6% had primary, pre-primary, or none and higher secondary education, respectively. Most of the participants (96.7%) had a monogamous marriage, whereas 3.2% belonged to polygamous marriage. Regarding the wealth index, most participants (21.0%) were from the poor quintile, and only 17.3 % were from the richest quartile. Most of the study participants (94.8 %) reported having poor media access, and only 0.2% reported good access to media.

**Table 1:** Demographic characteristics of Bangladeshi women of reproductive age (15-49) (n=47692)

| Characteristics | Frequency (f) | Percentage (%) |
| --- | --- | --- |
| **Age group (n=47692)** |  |  |
| 15-24 years | 10251 | 21.5 □ % |
| 25-34 years | 17542 | 36.8 □ % |
| 35-49 years | 19899 | 41.7 □ % |
|  | Mean ± SD | 32.6 ± 8.5 |
| **Residence** |  |  |
| Urban | 9460 | 19.8 □ % |
| Rural | 38232 | 80.2 □ % |
| **Educational level of the women** |  |  |
| Pre-primary or none | 8822 | 18.5 □ % |
| Primary | 12388 | 26.0 □ % |
| Secondary | 20484 | 43.0 □ % |
| Higher secondary or more | 5998 | 12.6 □ % |
| **Marital Structure (n=47665)** |  |  |
| Polygamous | 1543 | 3.2 □ % |
| Monogamous | 46122 | 96.7 □ % |
| **Wealth index quintile** |  |  |
| Poorest | 9847 | 20.6 □ % |
| Poor | 10022 | 21.0 □ % |
| Middle | 9954 | 20.9 □ % |
| Rich | 9600 | 20.1 □ % |
| Richest | 8269 | 17.3 □ % |
| **Media access (n=47672)** |  |  |
| Poor access | 45214 | 94.8 □ % |
| Moderate access | 2340 | 4.9 □ % |
| Good access | 118 | 0.2 □ % |
| **Tolerance towards DV** |  |  |
| No tolerance | 34578 | 72.5 □ % |
| Low tolerance | 9723 | 20.4 □ % |
| High tolerance | 3391 | 7.1 □ % |
| **Method of contraception** |  |  |
| No | 15962 | 33.5 □ % |
| Yes | 31730 | 66.5 □ % |

DV: Domestic violence.

**Figure 1:**
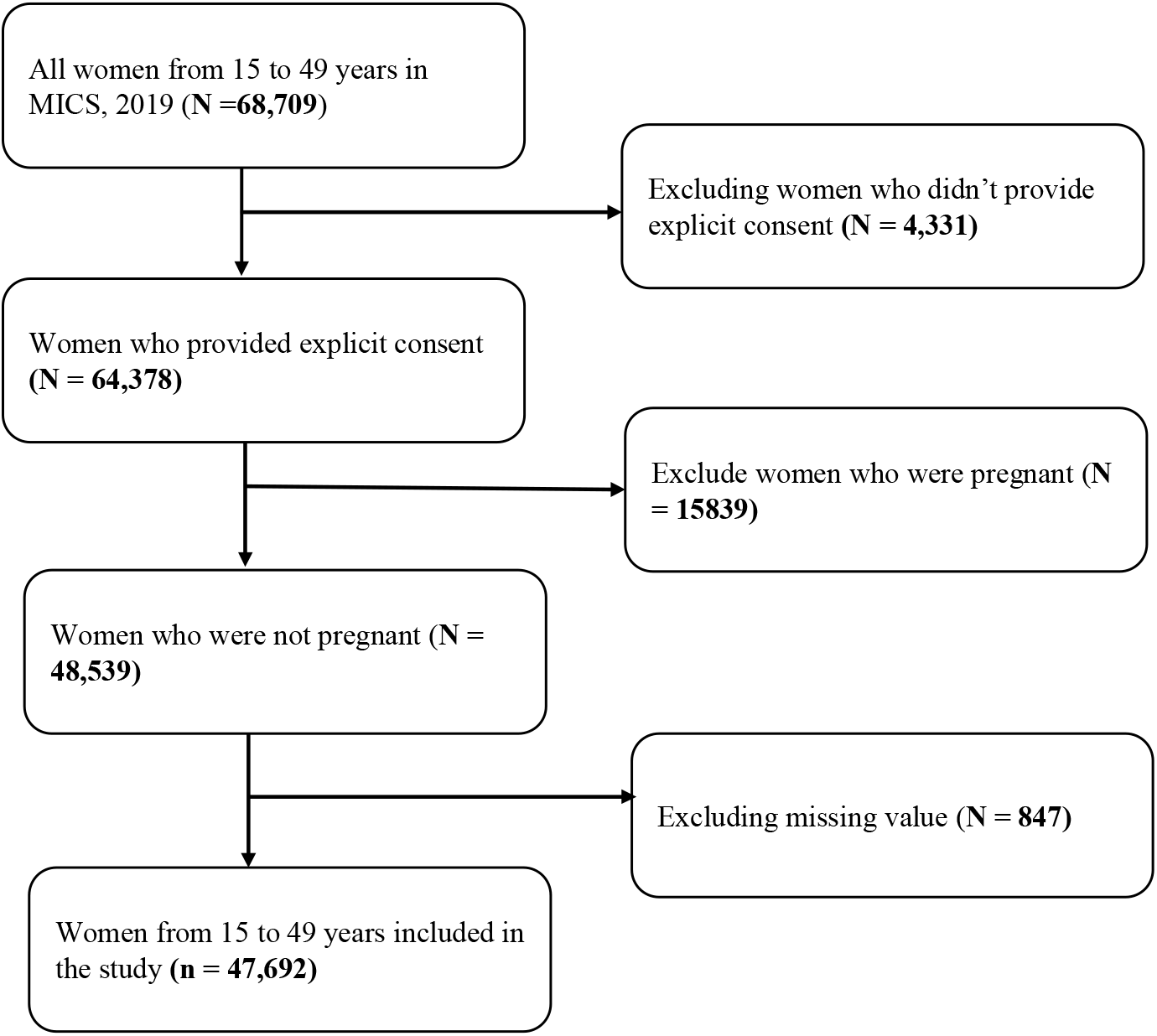
Selection of women of reproductive age (15-49) from the MICS 2019, Bangladesh

Regarding tolerance towards domestic violence, most participants (72.5%) had no tolerance regarding reasons for domestic violence. However, 7.1% of participants showed high tolerance. Regarding contraceptive use, most participants (66.5%) used contraceptive methods, and 33.5% did not.

A chi-square test of significance was used to determine the association between independent variables and methods of contraception use by the study participants. It was found that methods of contraception use was significantly associated with age group (p<0.001), residence (p<0.015), educational level of the women (p<0.001), marital structure (p<0.001), wealth index quintile (p<0.001), and tolerance towards DV (p<0.001) (Table 2).

**Table 2:**
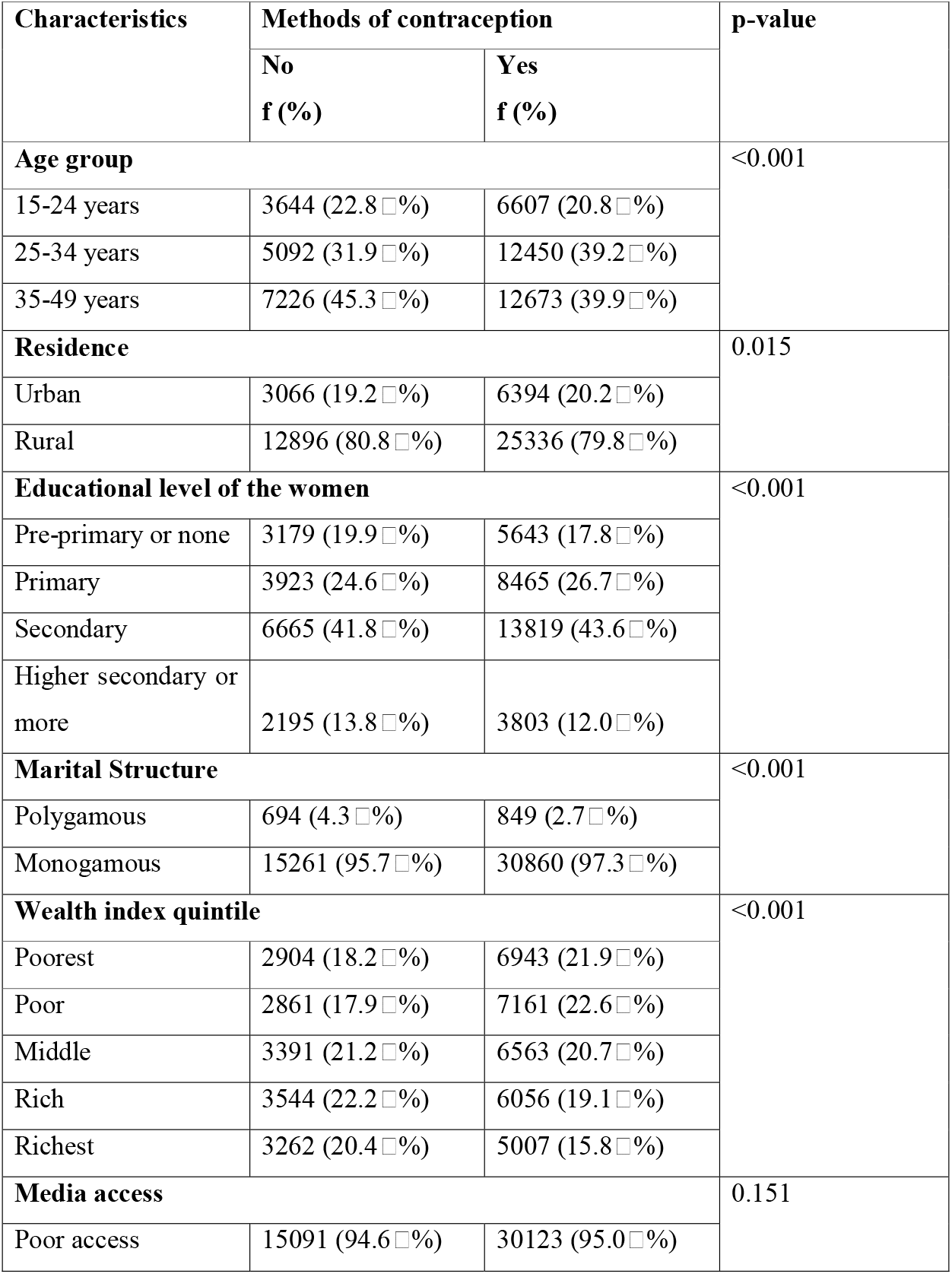
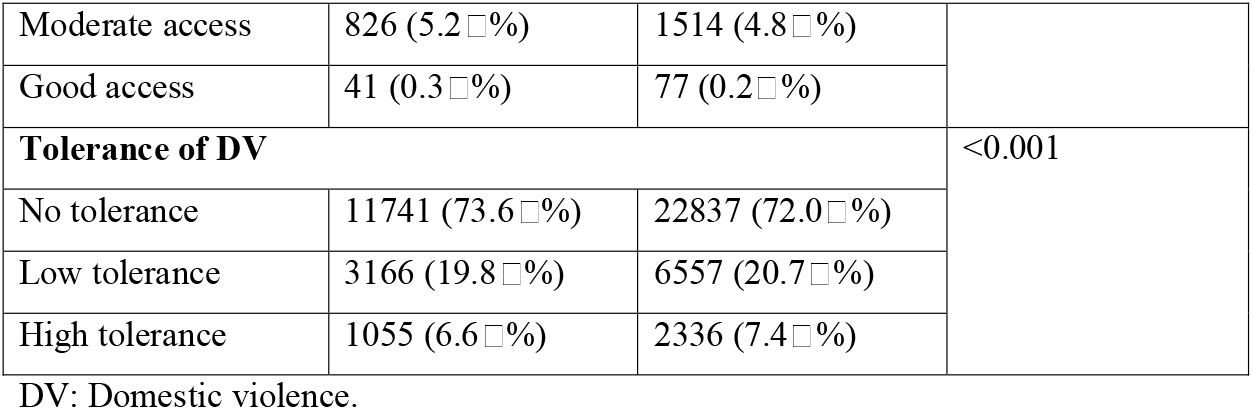
Association between Contraceptive use and important characteristics.

Using binary logistic regression, a multivariate analysis of all factors’ significance at 95% CI was conducted. It was found that age group, area, educational level of the women, residence, and educational level of the women, marital Structure, wealth index quintile, and tolerance towards DV were significantly associated with the use of CP.

The odds of CP use were 1.38 times higher among women in the age group 25-34 years compared to those in the age group of 15-24 years (AOR=1.382; 95% CI: 1.31-1.457, p-value < 0.001). Regarding residence, women living in rural areas had 0.747 (95% CI: 0.708-0.788, p-value <0.001) times lower odds of using CP than women of urban areas. In respect to educational level, those who belonged to the primary education, secondary education, and higher secondary or more education had AOR of using CP 1.212 (95% CI: 1.142-1.286; p-value <0.001), 1.264 (95% CI: 1.19-1.342; p-value <0.001) and 1.215 (95% CI: 1.122-1.316; p-value <0.001) respectively, compared to the group pre-primary or none. Women in polygamous marriages had lower odds of using CP than those in monogamous relationships (AOR=0.581; CI: 0.523-0.644; p-value <0.001). Those who belonged to the middle, rich, and richest wealth index quintile had odds of 0.77 (CI: 0.724-0.819; p-value <0.001); 0.651(CI: 0.611-0.694; p-value <0.001) and 0.521 (CI: 0.484-.0.56; p-value <0.001) respectively compared to poorest quintile. Regarding tolerance towards DV, women with high tolerance had lower odds of (AOR: 0.017; CI: 1.017-1.189; p-value<0.017) compared to those who had no tolerance. (Table 3).

**Table 3:**
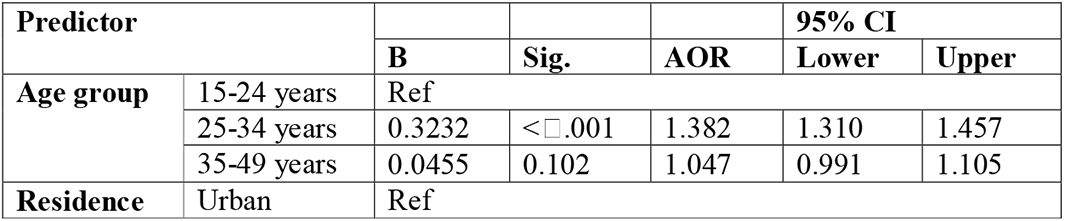
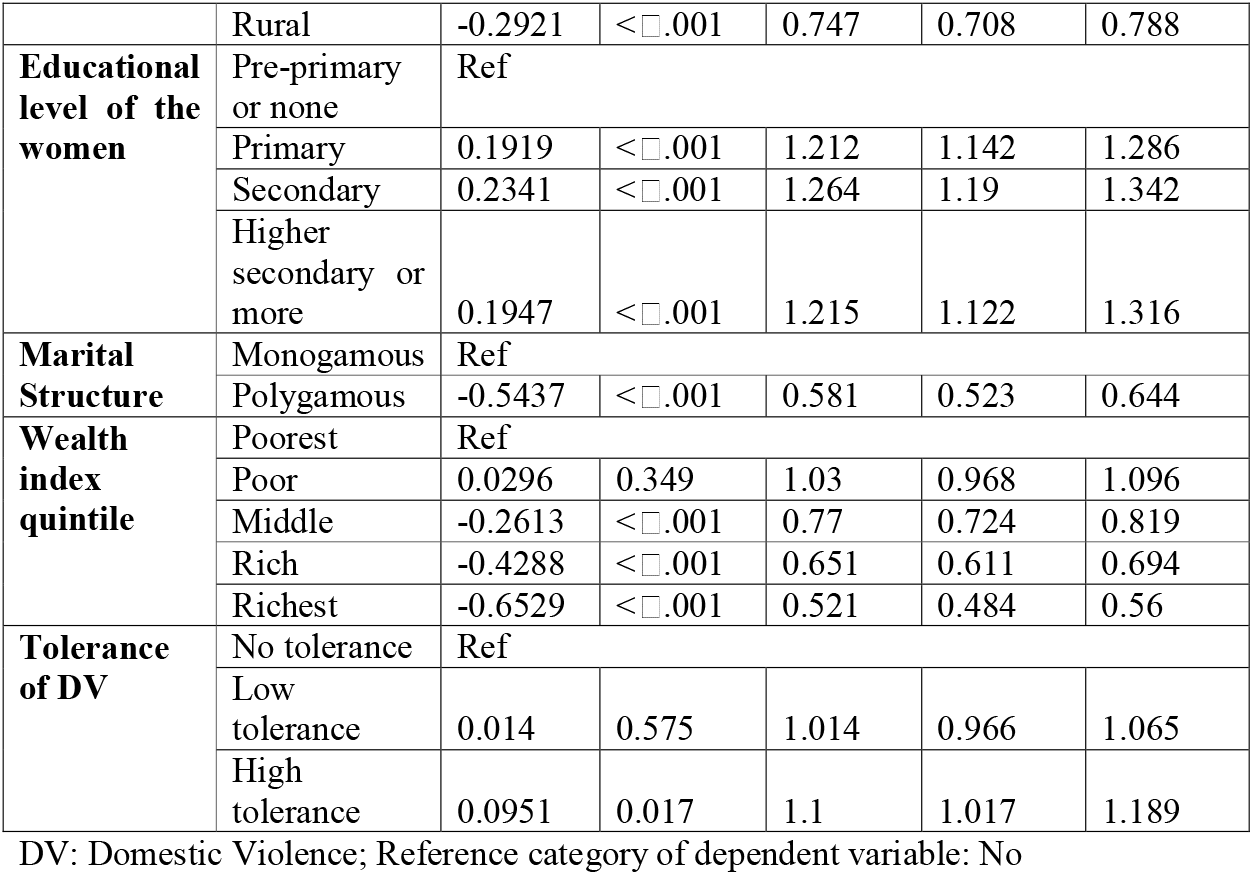
Binomial Multivariate logistic regression analysis of contraceptive use and significant independent variables.

## Discussion

The present study was conducted with data derived from MICS 2019, Bangladesh. The study aimed to investigate the use of contraception among Bangladeshi women of reproductive age (15-49 years) and its association with tolerance to domestic violence and other factors. This study found that the overall rate of contraceptive use was 66.5%, which collaborates with another study conducted in Bangladesh, which reports that 66.8% of participants used contraceptives.^26^

In the current study, women in the age group of 25-34 years were more likely to use contraception than women in the age group of 15-24 years. It is likely because women aged 25-34 years are more likely to have already experienced pregnancy and likely to have high awareness regarding family planning. Our finding has exact similarity with another Bangladeshi study conducted on contraceptive use in employed and unemployed women.^27^

The current study also suggests that women who live in rural areas are less likely to use contraception than women who live in urban areas. Women from urban areas usually have more self-autonomy and awareness regarding contraceptive use than women from rural areas. Moreover, they have better access to healthcare and family planning. Similar findings were found in one study conducted in 2020.^27^

The study found that women with higher level of education use contraceptive method more than women with lower levels of education. This finding collaborates with the claim of another research conducted in 2000.^28^ A higher level of education is known to increase awareness regarding the benefits of contraception and know-how regarding how to access them.

The current study indicates that women who are in monogamous marriages are more likely to use contraception than women who are in polygamous marriages. Women in polygamous marriages often suffer from an unstable household, lack of communication skills, and self-autonomy. A similar finding has been obtained in a study on polygamy with contraceptive use.^29^

The current study also reveals that women who belong to the wealthier quintiles were more likely to use contraception than those who belong to the poorer quintiles. It is because women in the wealthier quintiles have more access to healthcare services and information about contraception than their counterparts.

Women with a high tolerance towards domestic violence use less contraceptive methods than women without tolerance of domestic violence. This is because women with high tolerance towards DV usually suffer more atrocities of DV, making them vulnerable and unable to demand contraceptive access. A similar finding has been obtained from a study conducted in seven countries in West and central Africa.^2^

The study had several strengths and limitations. It utilized data from the MICS 2019 Bangladesh survey, a nationally representative survey. Also, the sample size used in this study was large. It also collaborates findings from several studies conducted in Bangladesh, adding to the existing knowledge. However, the study’s cross-sectional design renders it unable to establish causality. Moreover, as the study uses self-reported data, the possibility of recall bias must be addressed.

## Conclusion

The study’s findings have implications for the development of programs and policies to promote the use of contraception in Bangladesh. The findings suggest that programs and policies should focus on increasing access to contraception for women regardless of their age, educational level, household area, wealth index, marital status, and tolerance towards domestic violence. In addition, the current study findings suggest that programs and policies should focus on rural areas and target vulnerable populations like women with moderate or high tolerance towards DV. Other barriers like fear of their partners, fear of expressing the desire to use contraception, and lack of access to health care services may also hamper the rate of contraceptive use. Addressing these barriers, programs, and policies can help increase the use of contraception and improve the reproductive health of women in Bangladesh.

## Data Availability

Data will be shared by first author upon reasonable request.

## Acknowledgement

The authors are indebted UNICEF for providing us with access to the MICS 2019, Bangladesh dataset.

